# Population-Level Mortality Rates from Novel Coronavirus (COVID-19) in South Korea

**DOI:** 10.1101/2020.03.23.20041814

**Authors:** Samir Soneji, Hiram Beltrán-Sánchez, JaeWon Yang, Caroline Mann

## Abstract

**Background:** South Korea was among the first countries to report a case of the novel coronavirus (COVID-19) outside of China. As of 22 March, 2020, South Korea reported 8897 confirmed cases of and 104 deaths from COVID-19.

**Methods:** We collected the number of laboratory-confirmed cases and deaths in South Korea from the World Health Organization (as of 21 March, 2020) and case distribution and fatality rates by age from the Korean Center for Disease Control and Prevention (as of 22 March, 2020). We estimated population-level mortality rates by fitting a negative binomial regression model with the number of deaths as the outcome and population by age as an offset. We then calculated the age-standardized death rate (ASDR) based on the current COVID-19 figures and for alternative scenarios of increased prevalence.

**Findings:** The COVID-19 population-level mortality rate (per 100,000 person-years) increased with age: from 0.1 deaths among 30-39 year olds to 9.5 deaths among ≥80 year olds. The ASDR (per 100,000 person-years) was 0.8 deaths. The ASDR would increase to 52.0 deaths at a 1% prevalence (becoming the third leading cause of death) and 155.9 deaths at 3% prevalence (becoming the leading cause of death).

**Interpretation:** Currently, the population-level mortality burden of COVID-19 in South Korea, as measured by the ASDR, was relatively low compared to other causes of death partly due to the low prevalence of COVID-19. If the prevalence increases from another outbreak, the mortality burden could increase substantially and surpass other leading causes.

**Funding:** Grant P2C-HD041022, California Center for Population Research, University of California, Los Angeles (US NIH).

## INTRODUCTION

The first case of the novel coronavirus (COVID-19) in South Korea occurred in late January 2020, approximately two months after the first case globally occurred in the Hubei province of China.^1^ The number of cases quickly increased after an infected 61-year old woman attended religious services with over one thousand other people.^2^ Approximately one month after this first case and three days after the first death, South Korean President Moon Jae-In raised the national alert level to its highest possible level and instituted voluntary lockdown of affected cities and provinces.^3^ In addition to these precautionary measures, South Korea implemented widespread screening and has currently tested more people per capita than any other country.^4^

Case fatality rates are the most common measure used to assess the mortality burden of COVID-19. These rates are computed as the ratio of deaths due to COVID-19 to the number of people infected by the disease. However, comparison of COVID-19 case fatality rates across countries and historic pandemics is problematic because the *true* number of cases may far exceed the reported number of cases (i.e., the denominator of the case fatality rate) in most countries due to a lack of testing.^5^ Even in countries with widespread testing, the mortality burden of COVID-19 is more appropriately measured by age-specific mortality rates. In contrast to case fatality rates, mortality rates are computed by dividing deaths due to COVID-19 and the exposure measured in person-years lived (i.e., the entire population is at risk of infection and death from COVID-19). In addition, while case fatality rates provide a good assessment of the likelihood of death conditional on contracting the disease, mortality rates provide an overall risk of death for the entire population. These mortality rates can then be summarized into the age-standardized death rate (ASDR), which accounts for differences in the age distribution of populations and allows appropriate comparison across countries. The age distribution is an important factor because COVID-19 is more fatal among older adults and countries differ considerably in their proportion of older adults (e.g., 16% of the population of South Korea is aged ≥65 years compared to 12% in China and 23% in Italy).^6^

We address this research gap by utilizing standard demographic and epidemiological principles to estimate the population-level mortality rate for COVID-19 in South Korea. We focus on South Korea because it was among the first countries to report a case of COVID-19 and make public its case distribution and case fatality rates by age. This paper makes three important contributions. First, we estimate population-level mortality rates from COVID-19 by age. Second, we also consider alternative scenarios in which the prevalence of COVID-19 increased and compare the mortality burden of the observed pandemic and these alternative scenarios with leading causes of death in South Korea. Finally, we make available easy-to-use software for public health researchers, government officials, and international health organizations to input case distributions by age, case fatality rates by age, and the total number of cases (either laboratory confirmed or alternative scenarios) to estimate the mortality burden of COVID-19 for other countries.

## METHODS

### Data

We collected the number of laboratory-confirmed cases and total number of deaths in South Korea from 61 official World Health Organization (WHO) situation reports from 21 January 2020 to 21 March 2020 and supplemented with total cases and death from the South Korean Centers for Disease and Prevention (CDC) on 22 March 2020.^7^ We also utilized data from the South Korean CDC, which published the case distribution and case fatality rates in 10-year age groups for the 8897 cases that had been confirmed as of 22 March 2020.^8^ The case distribution equaled the number of cases in each age group divided by the total number of cases. The case fatality rate equaled the number of deaths in each age group divided by the number of cases in each age group. Finally, we utilized population counts by age group for South Korea in 2020 projected by the United Nations Department of Economic and Social Affairs.^9^

### Analysis

First, we began with the total number of COVID-19 cases and deaths to date for South Korea reported in WHO Situation Report Number 61 (21 March 2020). We then added the additional number of cases and deaths reported by the South Korean CDC on 22 March 2020. Thus, all data was current as of 22 March 2020. Second, we calculated the number of confirmed cases for each age group by proportionally allocating the total number of confirmed cases according to the case distribution by age group. Third, we calculated the total number of deaths by multiplying the number of confirmed cases and the case fatality rate for each age group. Fourth, we fit a negative binomial regression model on the mortality rates with the number of deaths as the outcome and the population by age as an offset. We then predicted mortality rates from the regression model. The mortality rates equaled the number of deaths divided by the number of person-years lived over the 63-day exposure period (time from first case on 20 January 2020 to 22 March 2020) for each age group. To convert mortality rates to an annual period, we multiplied them by the fraction of the year 2020 represented by the exposure period (i.e., 366 days/63 days). Sixth, we calculated the age-standardized death rates (ASDR) for COVID-19, where the age standard was based on the 2016 population in South Korea. We selected 2016 as the year of standardization because this is the most recent year for which the Vital Statistics Division of Statistics Korea published the ASDR of the leading causes of death (e.g., cancer and heart disease).^10^ See Appendix for step-by-step explanation of methods.

Finally we considered four alternative scenarios in which the prevalence of cases equaled 1%, 3%, 10%, and 25% of the population. These scenarios corresponded to 513,000; 1,538,000; 5,127,000; and 12,817,000 COVID-19 cases in South Korea, respectively (the population in 2020 is 51.3 million). We set 25% as the upper bound because this value was approximately the prevalence of the US population infected with influenza during the H1N1 influenza pandemic of 1918.^11^ For each scenario, we estimate the population-level mortality rate by age group and the ASDR. We compared the ASDR for COVID-19 in the observed pandemic and alternative scenarios with those of the 10 leading causes of death in South Korea.

## RESULTS

The first laboratory-confirmed case of COVID-19 in South Korea occurred on 21 January 2020 (Figure 1). The number of cases exceeded 100 on 20 February 2020 and 1000 on 26 February 2020. The first death occurred on 20 February 2020. The number of deaths exceeded 10 on 26 February 2020 and 50 on 9 March 2020. As of 22 March 2020 (63 days after the first confirmed case), South Korea reported 8897 cases and 104 deaths.

**Figure 1.**
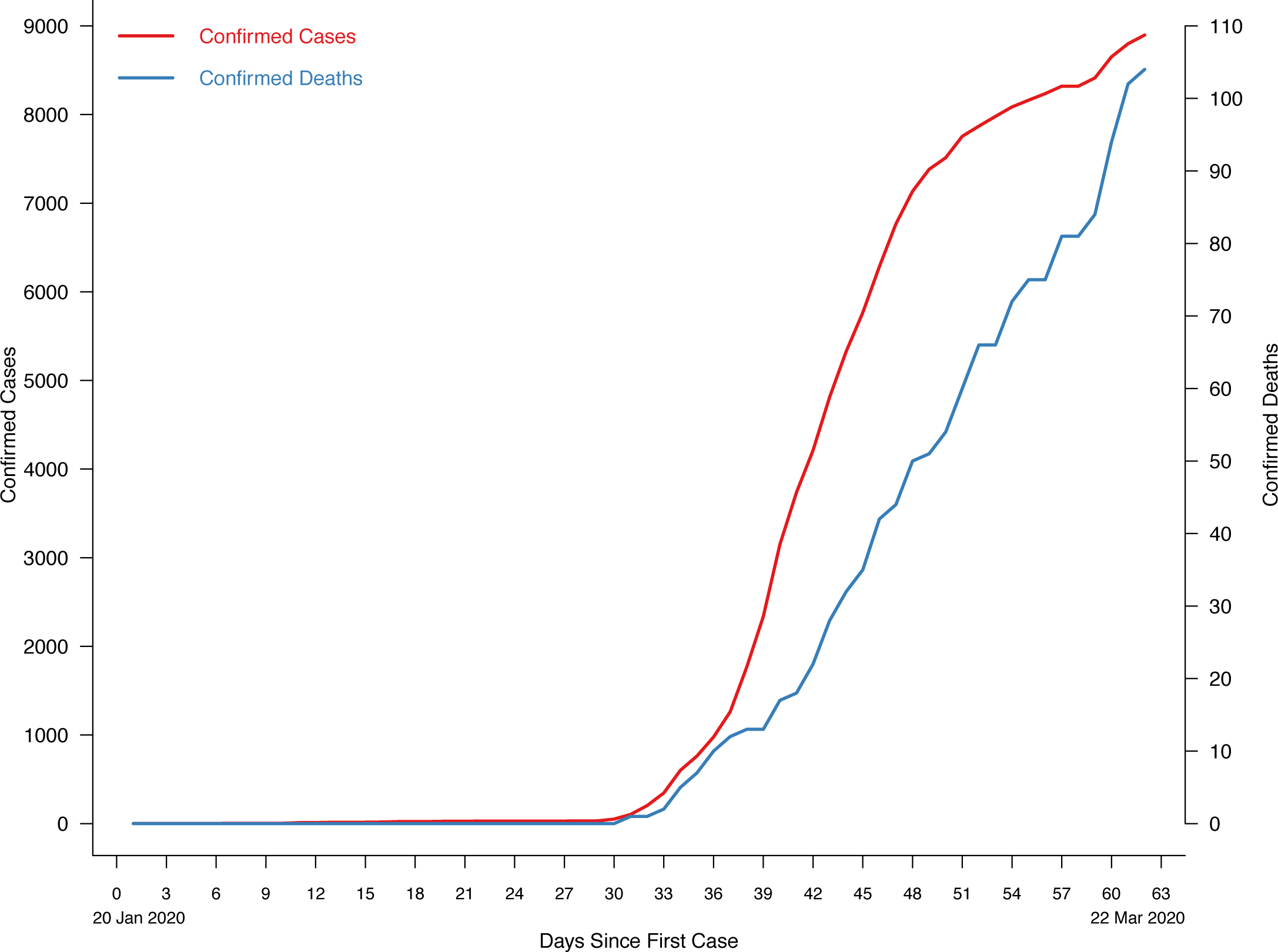
Number of Confirmed Cases of and Deaths from COVID-19, South Korea. Source: WHO Situation Reports, #1-#61 & Korean CDC

The distribution of cases by age group exhibited a bimodal distribution: 26.9% of cases occurred among individuals aged 20-29 years and 19.0% of cases among individuals aged 50-59 years (Table 1). The case fatality rate increased with age: from 0.11% among 30-39 year olds to 10.46% among ≥80 year olds. There were no reported deaths among those aged 29 years or younger. Our estimate of the population-level mortality rate also increased with age from 0.1 deaths per 100,000 person-years among 30-39 year olds (95% confidence interval [CI]: 0.0 to 0.2) to 12.8 deaths per 100,000 person-years among ≥80 year olds (95% CI: 9.6 to 17.2; Figure 2). The corresponding ASDR for COVID-19 was 1.0 deaths per 100,000 person-years (95% CI: 0.7 to 1.4).

**Table 1.** Population-Level Mortality Rate by Age Group for COVID-19, South Korea

| Age Group<br>(Years) | Population | # Cases | Proportion of<br>Population Infected | # Deaths | Mortality Rate<br>(Deaths per 100,000 Person-Years) |
| --- | --- | --- | --- | --- | --- |
| 0-9 | 4,153,813 | 101 | 0.00% | 0 | 0.00 |
| 10-19 | 4,753,258 | 460 | 0.01% | 0 | 0.00 |
| 20-29 | 6,716,294 | 2,396 | 0.04% | 0 | 0.00 |
| 30-39 | 7,079,839 | 909 | 0.01% | 1 | 0.01 |
| 40-49 | 8,218,844 | 1,221 | 0.01% | 1 | 0.01 |
| 50-59 | 8,476,699 | 1,691 | 0.02% | 7 | 0.08 |
| 60-69 | 6,453,706 | 1,132 | 0.02% | 17 | 0.26 |
| 70-79 | 3,560,646 | 595 | 0.02% | 37 | 1.04 |
| ≥80 | 1,856,084 | 392 | 0.02% | 41 | 2.21 |

Source: UN Population Projection, WHO Situation Report #57, and authors' calculations.

**Figure 2.**
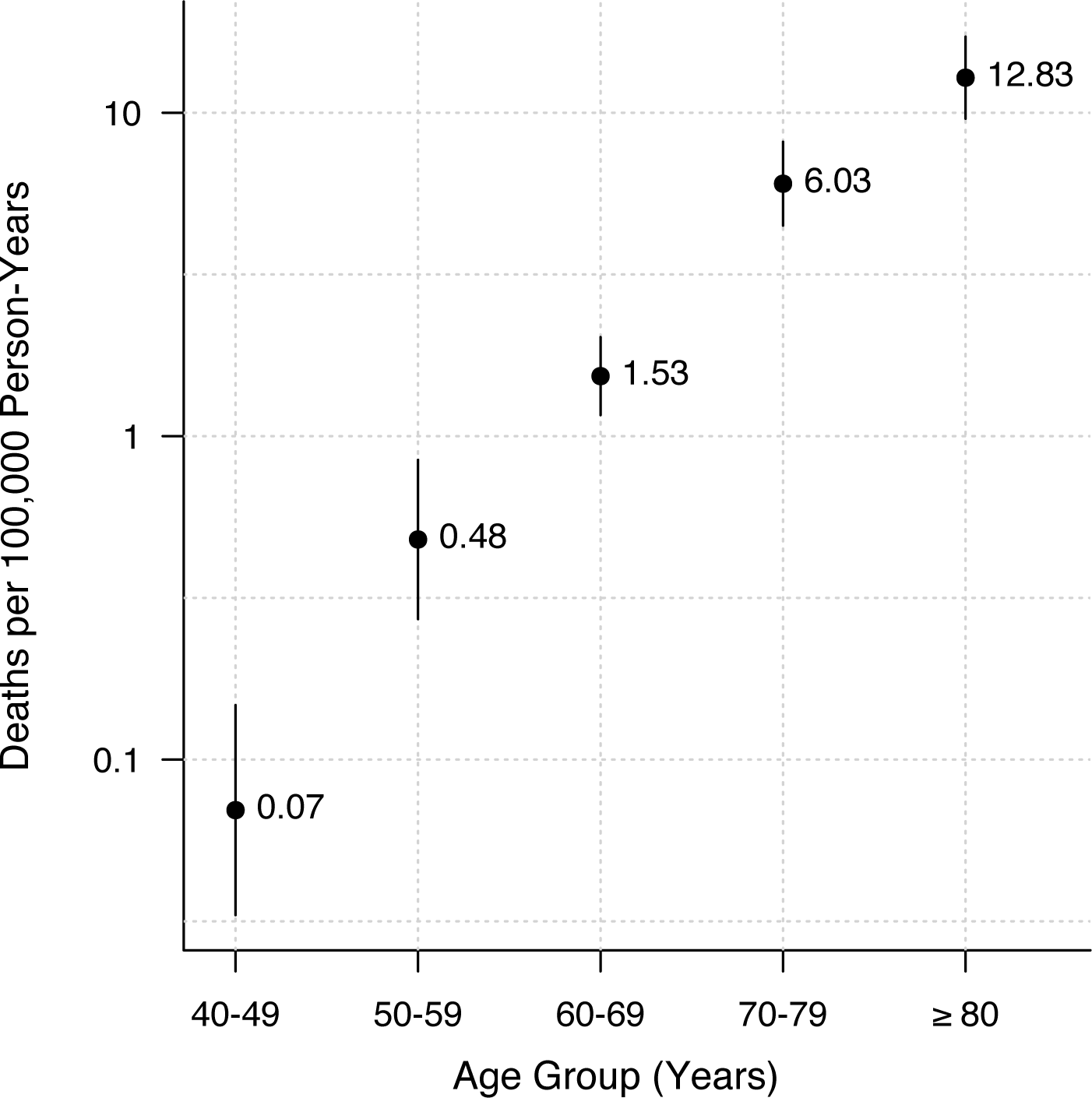
Population-Level Mortality Rates for COVID-19, South Korea. Source: authors’ calculation.

**Figure 3.**
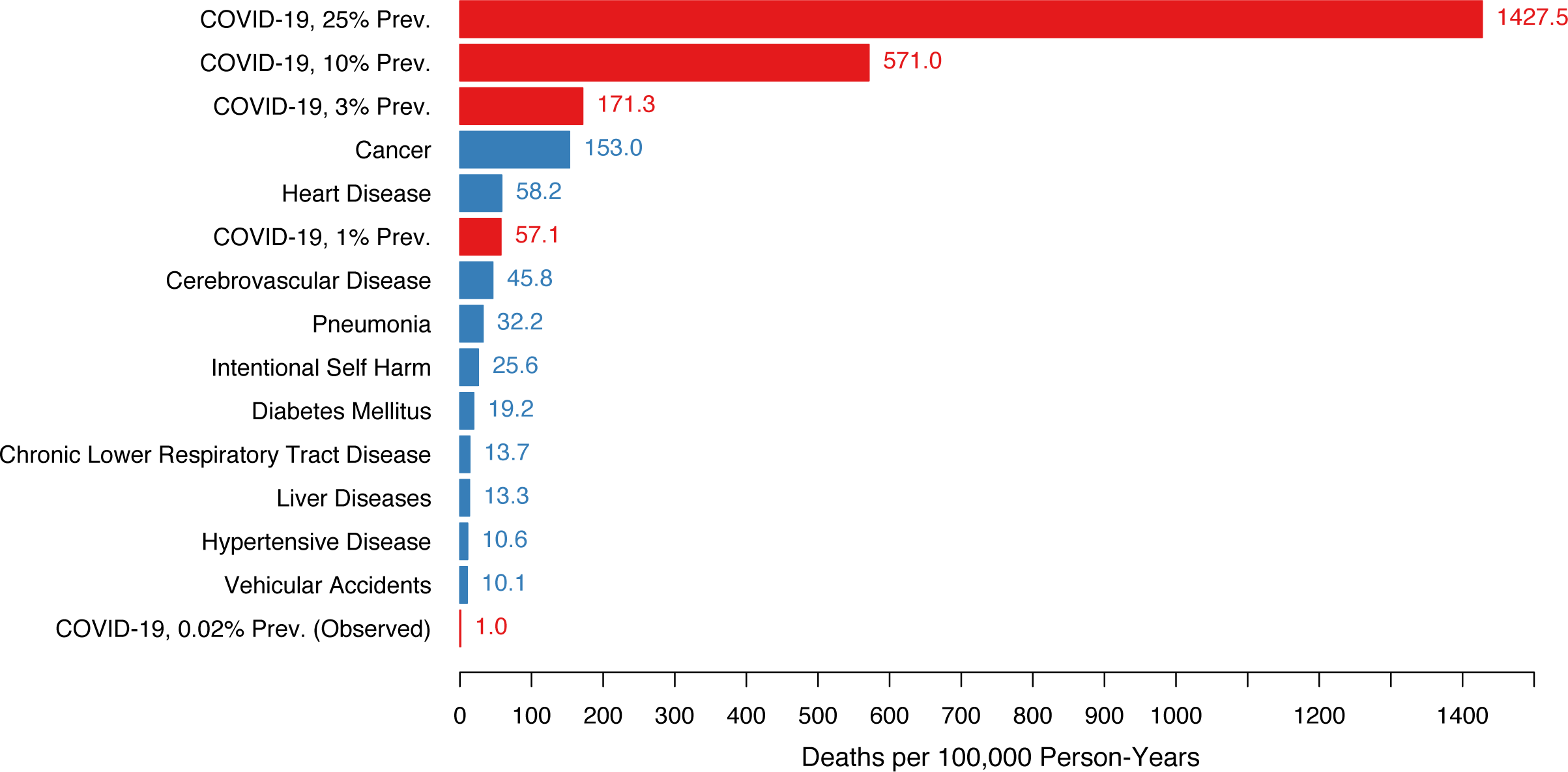
Age-Standardized Death Rate from Leading Causes of Death and COVID-19 (Observed and Alternative Scenarios), South Korea. Source: Statistics Korea and authors’ calculation.

Based on the current number of confirmed cases, case distribution, and case fatality rates, the ASDR for COVID-19 fell below the leading 10 causes of death in South Korea (Figure 2). However, relatively small increases in disease prevalence could quickly lead to a high mortality burden on the population. For example, if the prevalence of COVID-19 increased to 1% of the total population, the estimated ASDR for COVID-19 would equal 57.1 deaths per 100,000 person-years (95% CI: 54.7 to 59.6) and be only exceeded by heart disease and cancer. If the prevalence of COVID-19 equaled 3%, the estimated ASDR for COVID-19 would equal 171.3 deaths per 100,000 person-years (95% CI: 167.1 to 175.6) and approximately equal the ASDR for cancer—the leading cause of death in South Korea. Finally, if the prevalence of COVID-19 equaled 10%, the estimated ASDR for COVID-19 would equal 570.1 deaths per 100,000 person-years (95% CI: 563.3 to 578.8) and exceed the previous leading 10 causes of death combined.

## DISCUSSION

This study reports three central findings. First, the population-level mortality rate of COVID-19 in South Korea increased exponentially with age. Second, the population-level mortality burden, as measured by the ASDR, was relatively low compared to other causes of death because, in part, of the low prevalence of COVID-19. Third, the population-level mortality burden could increase substantially if the prevalence of COVID-19 increases. For example, at a prevalence level of 1%, COVID-19 would be the 3rd leading cause of death in South Korea, and at a prevalence level above 3%, it would surpass cancer as the leading cause of death.

The public, media, and government health agencies anxiously desire to know the possible mortality burden of COVID-19. However, the complex simulation models that estimate mortality outcomes require numerous assumptions and, perhaps more importantly, are inaccessible to most.^12–16^ In contrast, our approach utilizes public data published daily by the WHO and population-level clinical data that countries already collect.^17^ Our approach can be readily applied to other countries and will directly account for differences in the age distribution of populations, which is integral for calculating an accurate mortality burden as infected older adults may be more likely to die than infected middle-aged adults from COVID-19.

We note several limitations. First, the case distribution and case fatality rates on which we base our results were current as of 22 March 2020. However, the pandemic in South Korea, including the case distribution and case fatality rates could change rapidly.^2^ Another outbreak could overwhelm the capacity of the healthcare system and, consequently, case fatality rates may increase as limited healthcare resources are unable to accommodate the increasing number of COVID-19 patients.^18^ Second, our results are specific to South Korea, which was studied as their governmental agencies provided the most robust and updated case distribution and case fatality rate data by age. As a result, the ASDR of COVID-19 in South Korea, a nation which has been lauded for its widespread screening and early treatment, may differ from those of other countries.^19^ Our publicly available software allows researchers and public health officials to input case distributions by age, case fatality rates by age, and total number of cases (either laboratory confirmed or alternative scenarios) to estimate the mortality burden of COVID-19 for other countries. Third, the number of laboratory-confirmed cases could be less than the true number of cases, although South Korea has tested more individuals per capita to date than any other country. If the true number of cases exceeds the reported number of cases, the estimated number of deaths will increase and, thus, we may conservatively estimate the population-level mortality rate.

In conclusion, COVID-19 currently yields a relatively low mortality burden in South Korea compared to other causes of death because of its low prevalence in the population. If, however, the prevalence increases because of one or multiple future outbreaks or limited healthcare resources, the mortality burden could be substantially higher and exceed many leading causes of death.

## Data Availability

Full replication data for "Population-Level Mortality Rates from Novel Coronavirus (COVID-19) in South Korea" available on Harvard Dataverse.

https://doi.org/10.7910/DVN/SIJ1OC

## REFERENCES

1. Park WB, Kwon N-J, Choi S-J, et al. Virus Isolation from the First Patient with SARS-CoV-2 in Korea. J Korean Med Sci. 2020;35(7). doi:10.3346/jkms.2020.35.e84

2. Normile D. Coronavirus cases have dropped sharply in South Korea. What’s the secret to its success? Science. https://www.sciencemag.org/news/2020/03/coronavirus-cases-have-dropped-sharply-south-korea-whats-secret-its-success. Published March 17, 2020. Accessed March 19, 2020.

3. Cho J, Lee HK. South Korean president declares “war” on COVID-19 as deaths there reach 32. ABC News. https://abcnews.go.com/International/south-korean-president-declares-war-covid-19-deaths/story?id=69360757. Published March 3, 2020. Accessed March 19, 2020.

4. Bicker L. Coronavirus in South Korea: How “trace, test and treat” may be saving lives. BBC News. https://www.bbc.com/news/world-asia-51836898. Published March 12, 2020. Accessed March 19, 2020.

5. Thomas K. As Coronavirus Testing Increases, Some Labs Fear a Shortage of Other Supplies. The New York Times. https://www.nytimes.com/2020/03/11/health/coronavirus-testing-shortages.html. Published March 11, 2020. Accessed March 19, 2020.

6. CDCMMWR. Severe Outcomes Among Patients with Coronavirus Disease 2019 (COVID-19) — United States, February 12–March 16, 2020. MMWR Morb Mortal Wkly Rep. 2020;69. doi:10.15585/mmwr.mm6912e2

7. Coronavirus disease 2019 (COVID-19) Situation Report - 58. March 2020. https://www.who.int/docs/default-source/coronaviruse/situation-reports/20200318-sitrep-58-covid-19.pdf?sfvrsn=20876712_2. Accessed March 19, 2020.

8. The Updates on COVID-19 in Korea as of 22 March. Korean Center of Disease Control. http://www.cdc.go.kr. Published March 22, 2020. Accessed March 22, 2020.

9. World Population Prospects 2019. United Nations. https://population.un.org/wpp/Download/Standard/Population/. Published 2019. Accessed March 19, 2020.

10. Vital Statistics Division, Statistics Korea, Shin H-Y, Lee J-Y, et al. Cause-of-death statistics in 2016 in the Republic of Korea. J Korean Med Assoc. 2018;61(9):573. doi:10.5124/jkma.2018.61.9.573

11. Frost WH. The Epidemiology of Influenza. Public Health Rep 1896-1970. 1919;34(33):1823–1836. doi:10.2307/4575271

12. Li R, Pei S, Chen B, et al. Substantial undocumented infection facilitates the rapid dissemination of novel coronavirus (SARS-CoV2). Science. March 2020. doi:10.1126/science.abb3221

13. Kucharski AJ, Russell TW, Diamond C, et al. Early dynamics of transmission and control of COVID-19: a mathematical modelling study. Lancet Infect Dis. March 2020. doi:10.1016/S1473-3099(20)30144-4

14. Wu JT, Leung K, Leung GM. Nowcasting and forecasting the potential domestic and international spread of the 2019-nCoV outbreak originating in Wuhan, China: a modelling study. Lancet Lond Engl. 2020;395(10225):689–697. doi:10.1016/S0140-6736(20)30260-9

15. Ferguson NM, Laydon D, Nedjati-Gilani G, et al. Impact of Non-Pharmaceutical Interventions (NPIs) to Reduce COVID-19 Mortality and Healthcare Demand. Imperial College COVID-19 Response Team; 2020:20. https://www.imperial.ac.uk/media/imperial-college/medicine/sph/ide/gida-fellowships/Imperial-College-COVID19-NPI-modelling-16-03-2020.pdf.

16. Choi SC, Ki M. Estimating the reproductive number and the outbreak size of Novel Coronavirus disease (COVID-19) using mathematical model in Republic of Korea. Epidemiol Health. March 2020:e2020011. doi:10.4178/epih.e2020011

17. Zastrow M. South Korea is reporting intimate details of COVID-19 cases: has it helped? Nature. March 2020. doi:10.1038/d41586-020-00740-y

18. Fink S. Worst-Case Estimates for U.S. Coronavirus Deaths. New York Times. https://www.nytimes.com/2020/03/13/us/coronavirus-deaths-estimate.html. Published March 13, 2020. Accessed March 19, 2020.

19. Tharoor I. South Korea’s coronavirus success story underscores how the U.S. initially failed. The Washington Post. https://www.washingtonpost.com/world/2020/03/17/south-koreas-coronavirus-success-story-underscores-how-us-initially-failed/. Published March 17, 2020. Accessed March 19, 2020.

